# Exploring the Role of Microbiome in Susceptibility, Treatment Response and Outcome among Tuberculosis Patients from Pakistan; Study Protocol for a Prospective Cohort Study (Micro-STOP)

**DOI:** 10.1101/2021.11.10.21266176

**Authors:** Muhammad Shahzad, Zia Ul Haq, Simon C Andrews

**Author notes:** **Corresponding author** Dr. Muhammad Shahzad, BDS (Pakistan), PhD (UK), Associate Professor, Institute of Basic Medical Sciences, Khyber Medical University Peshawar, Pakistan.

## Abstract

**Introduction:** Tuberculosis (TB) caused by *Mycobacterium tuberculosis* is a common infectious disease associated with significant morbidity and mortality, especially in low and middle-income countries. Successful treatment of the disease requires prolonged intake (6 – 8 months) of multiple antibiotics with potentially detrimental consequences on the composition and functional potential of the human microbiome. The protocol described in the current study aims to identify microbiome (oral and gut) signatures associated with TB pathogenesis, treatment response and, outcome in humans.

**Methods and analysis:** Four hundred and fifty, newly diagnosed TB patients from three district levels (Peshawar, Mardan, Swat) TB diagnosis and treatment centers will be recruited in this non-interventional, prospective cohort study and will be followed and monitored until treatment completion. Demographic and dietary intake data, anthropometric measurement and blood, stool and salivary rinse samples will be collected at baseline, day 15, month-2 and end of the treatment. Additionally, we will recruit age (± 3 years) and sex-matched healthy controls (n=30). Blood sampling will allow monitoring of the immune response during the treatment, while salivary rinse and fecal samples will allow monitoring of dynamic changes in oral and gut microbiome diversity. Within this prospective cohort study, a nested case-control study design will be conducted to assess perturbations in oral and gut microbiome diversity (microbial dysbiosis) and immune response and compare between the patients groups (treatment success vs failure).

**Ethics and dissemination:** The study has received ethics approval from the Ethic Board of Khyber Medical University Peshawar, and administrative approval from Provincial TB Control Program of Khyber Pakhtunkhwa, Pakistan. The study results will be presented in national and international conferences and published in peer-reviewed journals.

**Trial registration number:** NCT04985994; pre-results

**Strengths and limitations of the study:**

- Prospective study design.
- Large sample size, several sampling time points, and samples taken before, during and after anti-tuberculosis treatment will ensure capture of an accurate picture of any emerging microbial dysbiosis and immune modulation; this will provide new understanding of how these two factors are mutually influenced and how they are impacted by treatment and outcome in TB patients.
- In addition to microbial signatures, collection of socio-demographic, anthropometric, nutrition and co-morbidities data will permit further subgroup analysis and meaningful results that will be shared with the scientific community.
- A potential weakness is that not all patients may be able to provide samples for all of the specified time points.
- Another weakness is the raised likelihood that some participants will withdraw since the study is carried out in resource poor communities

## Introduction

Tuberculosis (TB), caused by the bacterium *Mycobacterium tuberculosis* (Mtb), is a global public health issue and the second most common cause of death in humans by an infectious agent after HIV/AIDS (1). It is estimated that around one fourth (24.8%) of the world’s population has latent Mtb infection with no clinical symptoms and minimum chances of infecting others (2). Of these, 5–10% may develop active disease (characterized by clinical signs and symptoms of TB or microbiological confirmation of Mtb infection, or both) at some stage of their life, especially when the immune system is compromised due to ageing, HIV infections or malnutrition (3). According to a 2020 Global Tuberculosis Report by the World Health Organization (WHO), approximately 10.0 million new cases of TB were diagnosed in 2019, of which 1.2 million subsequently died (1). The TB problem is more severe in lower- and middle-income countries, including Pakistan. On a global scale, Pakistan ranks 5^th^ in terms of TB prevalence with an estimated 570,000 new cases reported in 2019 (1);. TB poses a significant burden on Pakistan’s already strained healthcare system and economy.

TB pathogenesis is a complex and dynamic process encompassing several stages that include disease susceptibility, infection, latent or active disease and treatment response or failure (4). Mbt has the ability to evade the immune system over prolonged time periods, even decades after exposure, in the form of latent TB (5). Once active, the disease is among the most difficult to treat infections requiring multidrug therapy for as long as 6–9 months. However, treatment failure is frequent (even where adherence to antibiotic therapy is maintained) at 15% for drug susceptible infections and 31% for drug resistant TB cases (6,7). Thus, in some population groups, mortality is around 12% (8). Furthermore, relapse and TB reactivation are also common public health issues (9). Although, a number of factors that modulate the risk of progression from one stage of the disease to another has been documented, the underlying biological modulators of the risk remains elusive especially with regard to the microbiome (10). Emerging evidence suggests that the microbiome is likely to play a critical role in TB pathogenesis as well as in treatment response and outcome, primarily due to multifaceted interactions between the pathogen, microbiome and host (4).

The human microbiome consists of various commensal consortia of microorganisms (bacteria, archaea, viruses and fungi) colonizing different habitats of the human body, such as the skin, gut and mucosal surfaces (11). Recent technological advances spurred by culture-independent metagenomic sequencing techniques coupled with ever declining costs of sequencing (a 200,000-fold decrease since 2001) (12) have greatly improved our understanding of the microbiome’s crucial role in human health and disease. It is believed that humans have co-evolved with their commensal microbes for mutual benefit (13). The human host provides the microbiota with a warm and nutrient-rich environment in which to reside. On the other hand, the microbiome plays an important role in stimulating the development of a functional immune system (both innate and adaptive immune system), which is crucial in maintaining host-microbe symbiosis. The microbiota of the gut provides essential nutritional factors (such as vitamins) and enhances the host’s ability to extract energy from food. Indeed, there is now considerable evidence suggesting that the human microbiome impacts, either directly or indirectly, the development of various host processes including circadian rhythmicity and nutritional, metabolic and immune responses (14–16). Furthermore, the microbiome can exert such effects not only locally but also across distant body sites via gut-lung, gut-brain and/or gut-liver axes. Thus, any perturbation in microbiome composition or functional potential may lead to pathogenic infections and altered immune responses, and can also play a crucial role in development of non-communicable diseases ranging from immune-mediated disease to intergenerational obesity and even cancer (16). Microbial dysbiosis at distinct body sites, such as the oral cavity and the gut, has also been reported in TB-associated comorbidities such as diabetes mellitus (17) and malnutrition (18). However, until now, very few studies have focused on the role of key microbiome communities in response to TB infection.

Microbial dysbiosis in the human gut during TB infection was first reported by Dubourg and colleagues (19). Using culture-dependent and independent methods, they found impoverished microbial communities residing in the gut of a single TB patient undergoing anti-tuberculosis treatment. However, the study included only one human subject (a 63 year old female) with multidrug resistant TB. The patient was also on broad-spectrum antibiotic treatment for the preceding four months and it is unclear to what degree the reduction in gut bacterial diversity was due to the antibiotic treatment, MDR tuberculosis, nutritional status or other co-morbid conditions. Another study also found significant changes in gut microbial diversity in a mouse Mtb infection model where 16S-rRNA-gene NGS revealed a rapid reduction in gut microbial diversity following infection. It was suggested that this effect arose as a consequence of immune signaling from lung to gut (20) through the so called “gut-lung axis” whereby alterations in one microbiome causes an adjusted immune response and altered composition of the other. A more recent study by Yongfei Hu *et al* (2019) found unique gut microbiome signatures and metabolic functions that were predictive of Mtb infection in TB patients (21). Similarly, oral anaerobic bacteria such as *Prevotella* have been found in abundance in the lower respiratory tract of HIV patients undergoing anti-retroviral therapy. Metabolic products from these bacterial population, such as butyrate and other short chain fatty acids, were associated with increased susceptibility to TB infection in these patients (22). On the other hand, *Helicobacter pylori* infection in latent TB patients is thought to be protective against progression of disease from latent to active TB in humans (23). All these findings may be of relevance to the understanding of the role of the microbiota in TB pathogenesis, recurrence and reactivation. However, until now, there is no strong evidence that indicates whether there is any significant interplay between the microbiome and immune system in humans that affects susceptibility to TB infection, TB progression and response during the course of anti-tuberculosis therapy.

Therefore, this study aims to dissect the relationship between the microbiota (oral and gut) and its interaction with the immune system during TB infection and anti-tuberculosis therapy in humans.

## Objectives

### Primary objective

The primary objective of the study is to explore the effect of TB infection and anti-tuberculosis therapy on oral and gut microbiome diversity and functional potential, and the immune response in newly diagnosed TB patients from Pakistan.

### Secondary objectives

1. To determine oral and gut microbiome diversity and functional potential at baseline and compare with healthy controls.
2. To assess the relationship between the oral and gut microbiome and socio-demographic characteristics and dietary intake in TB patients at baseline, before the start of anti-tuberculosis treatment.
3. To describe any occurrence of oral and gut microbial dysbiosis and its association with adverse reaction and treatment failure in TB patients.
4. To identify specific oral and gut enterotypes associated with adverse reaction and unfavorable treatment outcomes.

## Methods and analysis

### Study design and setting

The ‘Microbiome in Susceptibility, Treatment Response and Outcome among Tuberculosis Patients (Micro-STOP)’ study is designed as a population-based, prospective cohort study in newly diagnosed TB patients in three district-level (Peshawar, Mardan and Swat) TB diagnostic and treatment centers of Khyber Pakhtunkhwa province of Pakistan. These centers have been selected based on the highest TB prevalence in the year 2020. The participating sites approximately treat 2500 of TB patients annually.

The study will be conducted as part of an ongoing collaboration between Khyber Medical University Peshawar, Pakistan, TB Control Program Khyber Pakhtunkhwa and University of Reading, UK. The TB Control Program is the main body for developing and implementing policies and strategies for prevention and management of TB, and for governing TB diagnostics and treatment centers across the province. The TB Control Program is responsible for patient identification, clinical data and collection of study-related samples. Khyber Medical University is the only public sector medical university and center for healthcare research and education in the province. The University of Reading will analyze the data as explained in the methodology section below. The Micro-STOP study started in August 2021 and data collection is planned till August 2022 to achieve the required sample size.

### Sample size

Because of the strict inclusion and exclusion criteria, and absence of similar studies, statistical power cannot be calculated accurately. Therefore, we have followed a pragmatic approach for sample size calculation. Previously published literature suggest that for metagenomic studies, a sample size of 30 is sufficient to observe phenotypic heterogeneity at the molecular level (24). We are aiming to recruit 450 TB patients on the basis of 7% TB treatment failure rate in Pakistan (1), to achieve sufficient numbers to observe phenotypic differences in the corresponding microbiomes (treatment failure vs success). Furthermore, we will also recruit 30 healthy controls.

### Study population

Newly-diagnosed, untreated TB patients will be recruited from TB-treatment facilities in selected centers of Khyber Pakhtunkhwa province of Pakistan. Controls will be healthy subjects with no history of pulmonary TB, matched for sex and age (±3 years) with the TB patient group. They will be randomly selected from non-family neighbors of the patients. One person will be selected randomly from 5 – 7 proposed healthy controls.

*Inclusion Criteria*. Participants will be included in the study if they are:

1. Diagnosed with pulmonary TB after detailed history collection, clinical examination and laboratory assessment (sputum culture positive).
2. Aged 18 - 65 years.
3. Willing to participate in the study.
4. Healthy controls are those who are free of TB symptoms, healthy on physical examination and provide a negative sputum culture result.

*Exclusion criteria*. Participants will be excluded if they are:

1. Already on anti-TB treatment or were previously treated for TB.
2. Severely anemic (Hb < 10g/dL).
3. Having diarrhea or other major gastrointestinal disorders.
4. Using a medically prescribed diet or nutritional supplement.
5. Pregnant or lactating women.
6. Patients with liver or renal dysfunction, or having any other chronic disease condition.
7. Extra pulmonary and drug resistance TB patients.

### Study and sampling procedure

An overview of the overall study flow and conduct is presented in Fig. 1. Patients will be recruited daily by respective site coordinators who are clinical experts in TB diagnosis and treatment provision.

**Figure 1:**
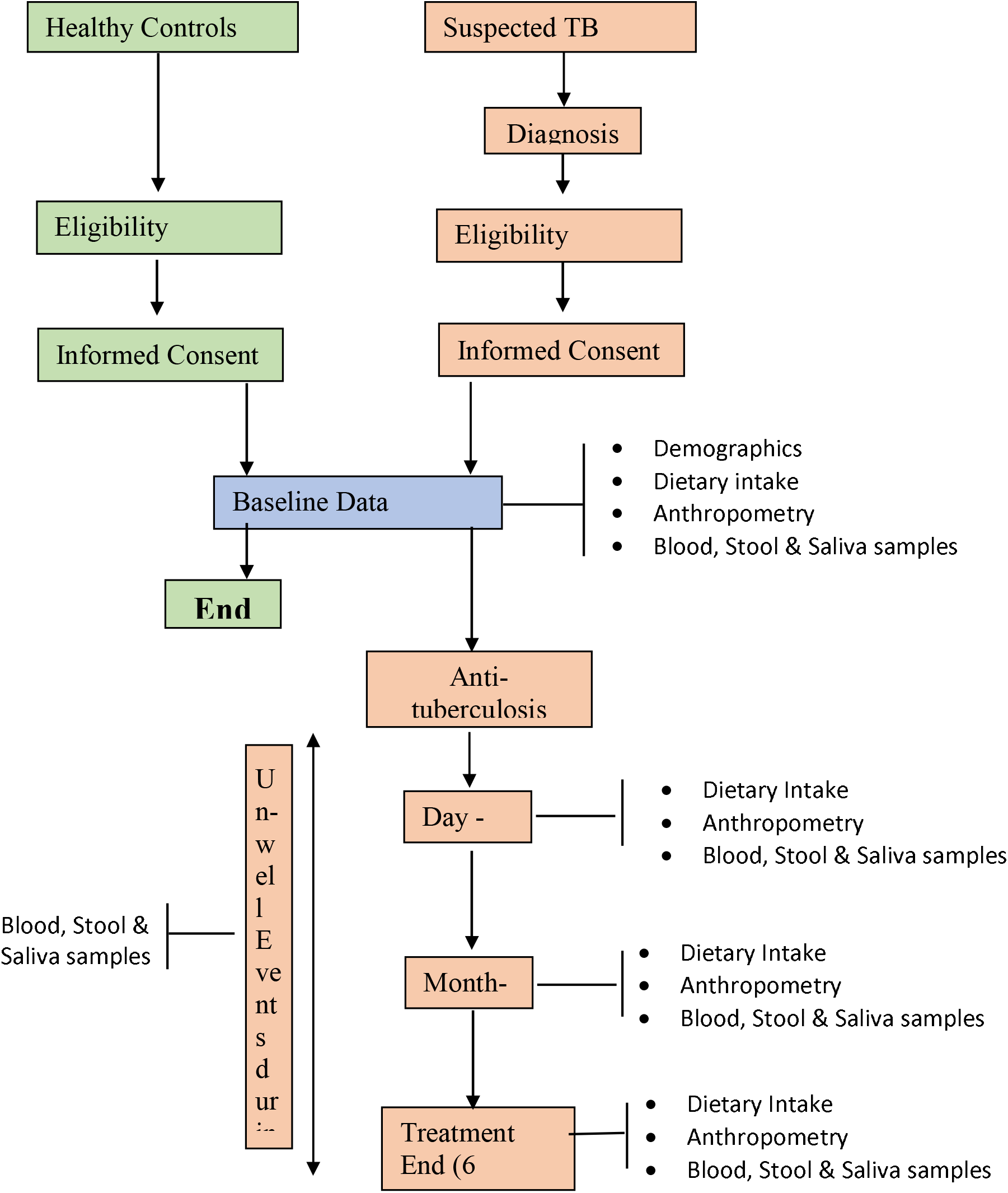
Flowchart of the study.

All patients with clinical or radiological signs suggestive of TB will undergo sputum smear microscopy and Xpert® MTB/RIF assay to confirm diagnosis of TB following the National TB Control Program revised guidelines for the control of TB in Pakistan. Smear microscopy employing Ziehl-Neelsen (ZN) staining to identify acid-fast bacilli (AFB) in sputum smears is a fast, easy and cost effective method for diagnosis of pulmonary TB in low and middle income countries (25). Xpert® MTB/RIF assay (Cepheid, Sunnyvale, CA, USA) is a WHO recommended, PCR-based molecular diagnostic technique for simultaneous identification of Mtb and rifampin resistance (i.e. mutation of the *rpoB* gene as surrogate for multidrug resistance) in clinical samples (26). The clinicians will explain the sample collection procedure to the patients in local language. The patients will be asked to provide up to 5 mL of blood-free sputum in sterile, 50 mL plastic jars with screw caps. Zeil Neelson staining of sputum smears will be performed and reporting will be based on the American Thoracic Society guidelines (27) described in Table 1.

**Table 1:** Reporting criteria for AFB smear microscopy.

| No of bacilli | Report |
| --- | --- |
| No AFB/200 field | No AFB seen |
| 1-9 AFB/100 field | Scanty |
| 10-99 AFB/100 field | + positive |
| 1-9 AFB/field | ++ positive |
| 10-99 AFB/field | +++ positive |

All the patients meeting the eligibility criteria will be invited to participate in the study by trained research assistants. A participant information sheet containing all the study details and procedures in an easy to understand, local language (Pashto) will be provided to all the participants. The study procedures will also be explained to the participants verbally. Once agreed, the participants will be asked to sign a written informed consent form in their preferred language.

### Collection of socio-demographic and anthropometric data

Socio-demographic information of the participants such as name, age, gender, ethnicity, education, occupation and household income will be collected by research assistants using structured questionnaire. The data will be collected only at baseline before the start of anti-tuberculosis therapy. Table 2 gives an overview of study conduct and timeline. Anthropometric measurements including height and weight will also be recorded following standard methods. In order to record height, patients will be instructed to remove shoes and head coverings (cap, scarf, etc), stand comfortably against a wall-mounted stadiometer (Seca GmBH, Hamburg, Germany) with heels, buttocks, shoulder blades and back of the head touching the vertical back board (Frankfurt plane position). Height will be then measured to the nearest 0.1 cm. Similarly for weight measurement, the patients will be asked to remove shoes and any extra piece of cloth and jewelry. Weight will be recorded in kilograms to the nearest 0.1 kg using calibrated electronic scale (Seca GmBH, Hamburg, Germany). Body mass index (BMI) will be computed as the fraction of weight in kilograms to the squared height in meters (kg/m^2^). All the anthropometric data will be collected at all four time points by two fully trained research assistants at each site.

**Table 2:**
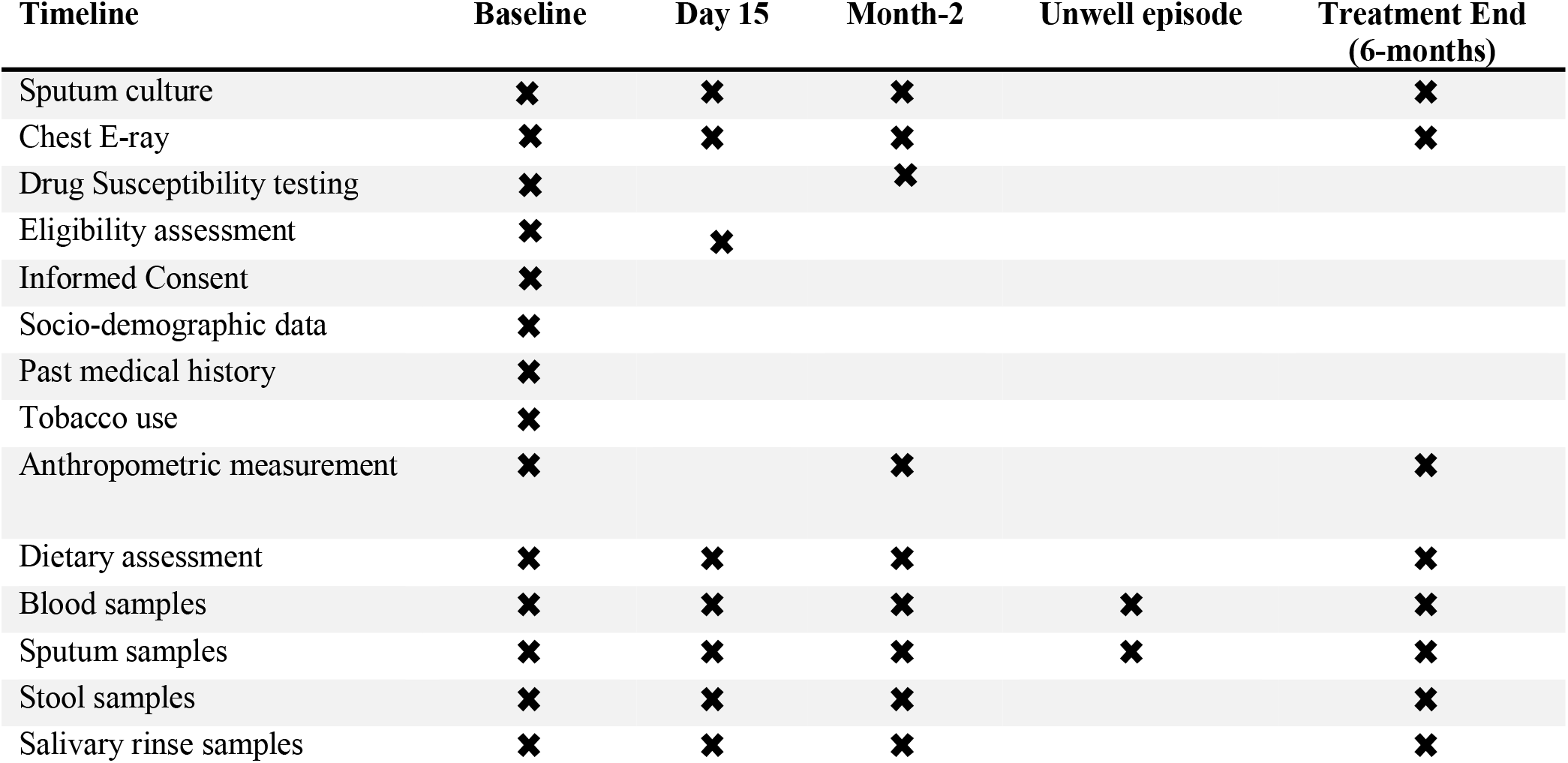
Study timeline and conduct.

### Dietary assessment

Because of the unavailability of a validated food frequency questionnaire for use in Pakistan, dietary intake of the participants will be assessed by trained nutritionist using the 24-h dietary recall method. The 24-h dietary recall will be conducted in the form of an in-depth interview using a standardized four stage protocol (28). The procedure typically requires around 20–30 minutes to complete. Efforts will be made to list all the food items and beverages consumed during the past 24-h period. This will include recording of information relating to all the food and beverages consumed, ingredients, cooking methods and brand names of commercial foods. The amount of each food or beverage consumed will be estimated in reference to common size containers (bowls, cups and glasses), standard measuring cups and spoons, two dimensional aids (photographs) and three dimensional food models. Dietary intake assessment will be performed at all four time points.

### Sample collection

#### Blood

Non-fasting, whole blood samples will be collected by a trained phlebotomist using butterfly needles and plastic vacutainers (BD Diagnostics, Switzerland). At least 5 mL of blood will be drawn from an antecubital vein into pre-chilled collection tubes containing silica clot activator. Whole blood will be sent to the local laboratory for complete blood count. For separation of serum, blood samples will be left on ice for 30 minutes followed by centrifugation. Aliquots of the serum samples (250 µL) will be prepared and stored at −80 °C till further processing in KMU main lab.

#### Stool

All participants will be asked to provide a stool sample at each of the four time points. For this purpose, stool samples collection kits will be provided which contain a collection pot, plastic bag, disposable gloves and an instruction sheet in their preferred language. Participants will be asked to pass an entire bowel movement into the plastic pot and close the lid when finished. After collection, the samples will be handed over to the research associate or delivered in their local study site (TB center) for further processing. The research associate will then transfer part of the stool samples to 10 mL screw top tubes (Bijoux tubes) with the help of the mini-spoon attached to the lid. Lids will be closed tightly, and tubes will then be placed in the plastic bag and transferred to the main laboratory maintaining the cold chain. In the laboratory, aliquots of the samples (200 mg) will be taken and stored at −80 °C for further processing.

#### Oral rinse

Oral rinse samples will be collected from all the patients at all the time points using standard methods (29). Before the sample collection, the patients will be instructed to avoid eating food, drinking fluids (especially flavored and carbonated drinks), chewing gum and using tobacco products for at least one hour before sample collection. They will be asked to swish (not gargle) 10□mL of sterile phosphate buffered saline (PBS) for 1□min, and expectorate the contents of the mouth into a 50□mL centrifuge tube. Collected samples will be vortexed (3□×□30□s), and each collected sample will be centrifuged at 14000 x g for 10□min at 4□°C to pelletize the corpuscular part and separate it from extracellular soluble components (supernatant). Resultant cell pellets will be frozen at −80□°C until use.

### Laboratory analysis

#### Hematological and biochemical parameters

Whole blood samples will be utilized for complete blood counts, and for hemoglobin, hematocrit, and mean corpuscular volume determination using an automated hematology analyzer (Sysmex XP-100, Singapore). Serum iron, ferritin, transferrin, C-reactive protein and alpha 1-acid glycoprotein (AGP) will be assayed using commercially available kits on an Abbott Architect ci8200 automated analyser (Abbott, Abbott Park, Illinois, USA) in the Mardan Medical Complex, Mardan, Pakistan. Micronutrient status of the patients will also be evaluated at all four time points. Vitamin A and D will be assessed through ELISA while plasma zinc and other minerals will be measured using the inductively coupled plasma-mass spectrometry facility at the University of Reading, UK.

#### Assessment of immune status

Immune status of the host will be assessed by full blood counts and measuring a panel of pro-inflammatory (IFN-γ, TNF-α and IL-6) and anti-inflammatory cytokine (IL-10, TGF-β1) levels in serum. Selection of these cytokines is based at the recently published study where significant differences in their levels were observed between patients with active TB and healthy controls (17). Serum concentration of cytokines will be measured by the sandwich ELISA technique following manufacturer instructions.

#### DNA extraction and metagenomic sequencing

Bacterial genomic DNA will be extracted from the oral rinse and fecal samples (200 mg) stored at −80 °C using a ZymoBIOMICS DNA Miniprep Kit (Zymo Research, Irvine, CA, USA) following the manufacturer’s instructions. Each sample will be quantified using a Denovix and quality-checked through agarose gel electrophoresis and PCR of the 16S rRNA gene target. To prepare the samples for sequencing, genomic DNA will be used to amplify the V1-V3 regions of the 16S rRNA gene using Golay barcoded primers through conventional PCR (30). The amplified samples will then be subjected to agarose gel electrophoresis to separate the DNA samples from primer dimers. Each DNA band will then be extracted and purified using a QIAquick gel extraction kit (Cat. no. 28705 QIAGEN® Germany). After purification, the concentration of each sample will be quantified using a Nanodrop. Concentrations of all samples will then be standardized to a minimum of 2.5 nmol. One µL of DNA from each of these samples will then be pooled together and sent for high throughput sequencing on the Illumina Miseq® (Illumina, San Diego, California, USA) platform at the Rehman Medical Institute, Peshawar following manufacturer instruction.

#### NGS data analysis

Sequence reads obtained for each sample will be processed for quality, sequence length, number of reads and removal of chimeras, which will be followed by organization into identified taxonomic units using QIIME software V.1.9.0 (31). Descriptive data will be presented and analyzed using appropriate statistical tests. For microbiome multivariate analysis, alpha and beta diversity (Observed, Chao1, Shannon and Simpson Index) will be compared between groups using the Phyloseque R software package and/or CLC Genomics workbench. For univariate analysis, microbiota populations will be compared between groups and significant differences selected with Bonferroni correction. We will seek to identify any correlations between the collected metadata and microbiome composition, and identify any gut/oral microbiota trends associated with TB.

#### Patients and public involvement

The community members or the patients were not directly involved in study design, conduct, and outcome measures. However, the protocol was reviewed and approved by Provincial TB control program of Khyber Pakhtunkhwa, Pakistan. Clinicians and program managers of TB control program have close liaison with the patients and act as bridge between the researchers and patients. They have substantial contribution in designing data collection questionnaires, informed consent and ensuring confidentiality. The study findings that report a positive impact on clinical practice will be reported in annual health meetings and communicated to the patients through the clinicians. However, individual data will not be reported back to the patients.

#### Ethical considerations

Ethics approval of the study has already been obtained from the Ethics Board of Khyber Medical University (DIR/KMU-EB/PR/000858). A participant information sheet will be provided to all the patients in their preferred language (Urdu, Pashto). The information sheet contains all the relevant information regarding the exact nature of the study, procedures, potential risks and benefits involved and whom to contact in case of emergency or when further information is required. The study procedures will also be explained verbally by clinical staff and research assistants and any queries will be answered. Once the participants have agreed to be included in the study, written informed consent will be obtained. The informed consent will be dated and signed by both the participant and the research assistant who presented the information to the participant. In cases where the participant is unable (illiterate) to sign the consent form, a thumb print will be obtained. The informed consent clearly indicates that the patient’s participation in the study is purely voluntary and he/she can withdraw from the study anytime without any reason and the decision will not affect the standard of treatment he/she receives.

#### Dissemination

We intend to publish the results of the study in international, peer reviewed journals as they become available. The data will also be presented in scientific conferences and seminars.

## Data Availability

All data produced in the present study are available upon reasonable request to the corresponding author

## Authors Contribution

Muhammad Shahzad and Simon C Andrews conceived the project idea and research methodology. Zia Ul Haq is overseeing recruitment of the study participants, samples and data collection. All the authors have read and approved the final version of this manuscript.

## References

1. World Health Organization. Global tuberculosis report 2020 [Internet]. Geneva, Switzerland; 2020. Available from: https://www.who.int/news-room/fact-sheets/detail/tuberculosis

2. Cohen A, Mathiasen VD, Schön T, Wejse C. The global prevalence of latent tuberculosis: a systematic review and meta-analysis. Eur Respir J. 2019 Jan 1 [cited 2021 Jul 3]; Available from: https://erj.ersjournals.com/content/early/2019/06/12/13993003.00655-2019

3. Jilani TN, Avula A, Zafar Gondal A, Siddiqui AH. Active Tuberculosis. In: StatPearls [Internet]. Treasure Island (FL): StatPearls Publishing; 2021 [cited 2021 Jul 3]. Available from: http://www.ncbi.nlm.nih.gov/books/NBK513246/

4. Naidoo CC, Nyawo GR, Wu BG, Walzl G, Warren RM, Segal LN, et al. The microbiome and tuberculosis: state of the art, potential applications, and defining the clinical research agenda. Lancet Respir Med. 2019 Oct 1;7(10):892–906.

5. Orme IM, Robinson RT, Cooper AM. The balance between protective and pathogenic immune responses in the TB-infected lung. Nat Immunol. 2015 Jan;16(1):57–63.

6. Karumbi J, Garner P. Directly observed therapy for treating tuberculosis. Cochrane Database Syst Rev. 2015 May 29;(5):CD003343.

7. Orenstein EW, Basu S, Shah NS, Andrews JR, Friedland GH, Moll AP, et al. Treatment outcomes among patients with multidrug-resistant tuberculosis: systematic review and meta-analysis. Lancet Infect Dis. 2009 Mar;9(3):153–61.

8. Shuldiner J, Leventhal A, Chemtob D, Mor Z. Mortality after anti-tuberculosis treatment completion: Results of long-term follow-up. Int J Tuberc Lung Dis. 2016 Jan 1;20:43–8.

9. Humbwavali JB, Trujillo NJ, Paim BS, Wolff FH, Barcellos NT. Sputum monitoring during tuberculosis treatment for predicting outcome. Lancet Infect Dis. 2011 Mar;11(3):160; author reply 160-161.

10. Barry CE, Boshoff HI, Dartois V, Dick T, Ehrt S, Flynn J, et al. The spectrum of latent tuberculosis: rethinking the biology and intervention strategies. Nat Rev Microbiol. 2009 Dec;7(12):845–55.

11. Gilbert JA, Blaser MJ, Caporaso JG, Jansson JK, Lynch SV, Knight R. Current understanding of the human microbiome. Nat Med. 2018 Apr;24(4):392–400.

12. The Cost of Sequencing a Human Genome [Internet]. Genome.gov. [cited 2021 Sep 29]. Available from: http://www.genome.gov/about-genomics/fact-sheets/Sequencing-Human-Genome-cost

13. Dethlefsen L, McFall-Ngai M, Relman DA. An ecological and evolutionary perspective on human–microbe mutualism and disease. Nature. 2007 Oct;449(7164):811–8.

14. Hacquard S, Garrido-Oter R, González A, Spaepen S, Ackermann G, Lebeis S, et al. Microbiota and Host Nutrition across Plant and Animal Kingdoms. Cell Host Microbe. 2015 May 13;17(5):603–16.

15. Lynch JB, Hsiao EY. Microbiomes as sources of emergent host phenotypes. Science. 2019 Sep 27;365(6460):1405–9.

16. Zheng D, Liwinski T, Elinav E. Interaction between microbiota and immunity in health and disease. Cell Res. 2020 Jun;30(6):492–506.

17. Qin J, Li Y, Cai Z, Shenghui L, Zhu J, Zhang F, et al. A metagenome-wide association study of gut microbiota in type 2 diabetes. Nature. 2012 Sep 26;490:55–60.

18. Million M, Diallo A, Raoult D. Gut microbiota and malnutrition. Microbiota Nutr. 2017 May 1;106:127–38.

19. Dubourg G, Lagier JC, Armougom F, Robert C, Hamad I, Brouqui P, et al. The gut microbiota of a patient with resistant tuberculosis is more comprehensively studied by culturomics than by metagenomics. Eur J Clin Microbiol Infect Dis. 2013 May 1;32(5):637–45.

20. Winglee K, Eloe-Fadrosh E, Gupta S, Guo H, Fraser C, Bishai W. Aerosol Mycobacterium tuberculosis Infection Causes Rapid Loss of Diversity in Gut Microbiota. PLOS ONE. 2014 May 12;9(5):e97048.

21. Hu Y, Feng Y, Wu J, Liu F, Zhang Z, Hao Y, et al. The Gut Microbiome Signatures Discriminate Healthy From Pulmonary Tuberculosis Patients. Front Cell Infect Microbiol. 2019;9:90.

22. Segal LN, Clemente JC, Li Y, Ruan C, Cao J, Danckers M, et al. Anaerobic Bacterial Fermentation Products Increase Tuberculosis Risk in Antiretroviral-Drug-Treated HIV Patients. Cell Host Microbe. 2017 Apr 12;21(4):530–537.e4.

23. Perry S, de Jong BC, Solnick JV, de la Luz Sanchez M, Yang S, Lin PL, et al. Infection with Helicobacter pylori is associated with protection against tuberculosis. PloS One. 2010 Jan 20;5(1):e8804.

24. Ardura MI, Banchereau R, Mejias A, Di Pucchio T, Glaser C, Allantaz F, et al. Enhanced monocyte response and decreased central memory T cells in children with invasive Staphylococcus aureus infections. PloS One. 2009;4(5):e5446.

25. Chandra TJ, Alan RR, Selvaraj R, Sharma YV. MODS ASSAY FOR RAPID DIAGNOSIS OF TUBERCULOSIS AMONG HIV TB CO INFECTED INDIVIDUALS IN A TERTIARY CARE HOSPITAL, ANDHRA PRADESH. Pak J Chest Med. 2014 20(4).

26. Boehme CC, Nabeta P, Hillemann D, Nicol MP, Shenai S, Krapp F, et al. Rapid Molecular Detection of Tuberculosis and Rifampin Resistance. N Engl J Med. 2010 Sep 9;363(11):1005–15.

27. Lewinsohn DM, Leonard MK, LoBue PA, Cohn DL, Daley CL, Desmond E, et al. Official American Thoracic Society/Infectious Diseases Society of America/Centers for Disease Control and Prevention Clinical Practice Guidelines: Diagnosis of Tuberculosis in Adults and Children. Clin Infect Dis. 2017 Jan 15;64(2):e1–33.

28. Shim J-S, Oh K, Kim HC. Dietary assessment methods in epidemiologic studies. Epidemiol Health. 2014 Jul 22;36:e2014009.

29. Woo JS, Lu DY. Procurement, Transportation, and Storage of Saliva, Buccal Swab, and Oral Wash Specimens. Methods Mol Biol Clifton NJ. 2019;1897:99–105.

30. Zheng W, Tsompana M, Ruscitto A, Sharma A, Genco R, Sun Y, et al. An accurate and efficient experimental approach for characterization of the complex oral microbiota. Microbiome. 2015 Oct 5;3:48.

31. Caporaso JG, Kuczynski J, Stombaugh J, Bittinger K, Bushman FD, Costello EK, et al. QIIME allows analysis of high-throughput community sequencing data. Nat Methods. 2010 May;7(5):335–6.

